# Opportunistic screening for coronary artery calcium deposition using chest radiographs – a multi-objective models with multi-modal data fusion

**DOI:** 10.1101/2024.01.10.23299699

**Authors:** Jiwoong Jeong, Chieh-Ju Chao, Reza Arsanjani, Chadi Ayoub, Steven J. Lester, Milagros Pereyra, Ebram F Said, Michael Roarke, Cecilia Tagle-Cornell, Laura M. Koepke, Yi-Lin Tsai, Chen Jung-Hsuan, Chun-Chin Chang, Juan M. Farina, Hari Trivedi, Bhavik N. Patel, Imon Banerjee

## Abstract

**Background:** To create an opportunistic screening strategy by multitask deep learning methods to stratify prediction for coronary artery calcium (CAC) and associated cardiovascular risk with frontal chest x-rays (CXR) and minimal data from electronic health records (EHR).

**Methods:** In this retrospective study, 2,121 patients with available computed tomography (CT) scans and corresponding CXR images were collected internally (Mayo Enterprise) with calculated CAC scores binned into 3 categories (0, 1-99, and 100+) as ground truths for model training. Results from the internal training were tested on multiple external datasets (domestic (EUH) and foreign (VGHTPE)) with significant racial and ethnic differences and classification performance was compared.

**Findings:** Classification performance between 0, 1-99, and 100+ CAC scores performed moderately on both the internal test and external datasets, reaching average f1-score of 0.66 for Mayo, 0.62 for EUH and 0.61 for VGHTPE. For the clinically relevant binary task of 0 vs 400+ CAC classification, the performance of our model on the internal test and external datasets reached an average AUCROC of 0.84.

**Interpretation:** The fusion model trained on CXR performed better (0.84 average AUROC on internal and external dataset) than existing state-of-the-art models on predicting CAC scores only on internal (0.73 AUROC), with robust performance on external datasets. Thus, our proposed model may be used as a robust, first-pass opportunistic screening method for cardiovascular risk from regular chest radiographs. For community use, trained model and the inference code can be downloaded with an academic open-source license from https://github.com/jeong-jasonji/MTL_CAC_classification.

**Funding:** The study was partially supported by National Institute of Health 1R01HL155410-01A1 award.

## Introduction

Coronary artery disease (CAD) remains a significant global health concern, contributing to substantial morbidity and mortality rates (1). As such, effective risk assessment and early intervention strategies are essential for reducing the burden of coronary artery calcification (CAC) (2). Computational tomography (CT) imaging has revolutionized the field through the use of CAC score (3). CAC score has been proven to be valuable for accurate risk stratification and guiding preventive interventions, such as statin therapy and aspirin administration, especially in asymptomatic individuals (4).

Despite the advantages of CT imaging in assessing CAC scores, certain inherent drawbacks limit its widespread application. Factors such as electrocardiogram (ECG)-gating requirements and the presence of arrhythmias pose challenges to obtaining reliable and accurate measurements until the development of qualitative ungated CAC score estimation (5). Moreover, conducting these CT exams often require significant technical, equipment, and clinical resources, which may not be operationally feasible at small healthcare facilities. While the practice is moving toward using non-gated CT scanning, a high-quality CAC study still requires multi-slice CT scanners and additional image processing and analysis by trained specialists for the quantification and interpretation of CAC score (6). Furthermore, CAC testing is not universally covered by insurance plans (7). While there is some evidence that CAC might be cost effective in patients with a family history of coronary disease (8), generally the measurement of CAC testing is not cost-effective for screening large populations to detect CAD in asymptomatic individuals.

In light of these limitations, there is growing clinical interest in exploring alternative methods that can simplify the assessment of CAC scores or detection of the potential plaque deposition on routine imaging modalities using opportunistic screening (9). One potential solution lies in the combination of regular chest radiographs/X-rays (CXR) and artificial intelligence (AI) technology. While CXR is not conventionally considered the primary modality for directly assessing coronary artery conditions, recent advancements in deep learning techniques have opened new possibilities. By leveraging AI algorithms, it becomes feasible to extract the calcification features present in CXR images, allowing for the opportunistic estimation of CAC scores. Compared to CT imaging, a CXR-based AI approach could offer several distinct advantages. Firstly, it significantly decreased ionizing radiation exposure (0.1 mSv versus 0.8-10.5 mSv), therefore reducing the associated health risks (10,11). This attribute is particularly crucial for repeated or serial screenings, enabling longitudinal monitoring of CAC scores without increasing patients’ radiation exposure, roughly nine CXR doses equaling one CT dose (12). Additionally, CXR is a widely available and cost-effective imaging modality, making it nearly universally accessible across various healthcare settings (13).

This study aims to investigate the potential of utilizing CXR and AI technologies to identify and stratify patients with coronary atherosclerosis, offering a low-radiation and cost-effective alternative for risk assessment. We hypothesize that by applying deep learning algorithms to CXR images, it is possible to extract the necessary calcification features and accurately identify high CAC category which can provide an efficient way of detecting asymptomatic individuals. Through this approach, we anticipate providing clinicians with a simpler and more accessible tool for risk stratification, ultimately facilitating timely interventions and improving patient outcomes related to CAD with cost-efficient imaging modality.

## Materials and Methods

### Internal cohort

We collected a retrospective internal cohort of 2,121 patients who had a coronary CT scan between 2012 - 2022 and corresponding CXR imaging exam with frontal view within a ± 1 year period of the CT exam day at the Mayo Clinic. If a patient underwent multiple radiography or CT examinations, all were included in the data set, with radiographs linked to the temporally closest calcium scoring CT (Table 1). Informed consent waiver and ethical approval was obtained from the Mayo Clinic Institutional Review Board (IRB protocol#22-006839). The 2/3 of the internal dataset was used for training and the 1/3 was split into validation and testing resulting in 1,424/356/341 patients, respectively. CAC scoring was performed on cardiac chest CT according to standard imaging acquisition and Agatston scoring methods (14). We extracted the CAC score from clinical radiology reports using simple regular expression (REGEX) and manually reviewed the extracted scores. After extraction, CAC scores were grouped into three bins based on accepted clinical cutoffs and literature defining prognosis by CAC category (15,16): ‘no CAC’ - 0, 1-99, and 100+. Additionally, patients with CAC 400+ were also identified to test the models’ performance in differentiating high vs. low-risk cases and compare to available literature (9).

**Table 1.**
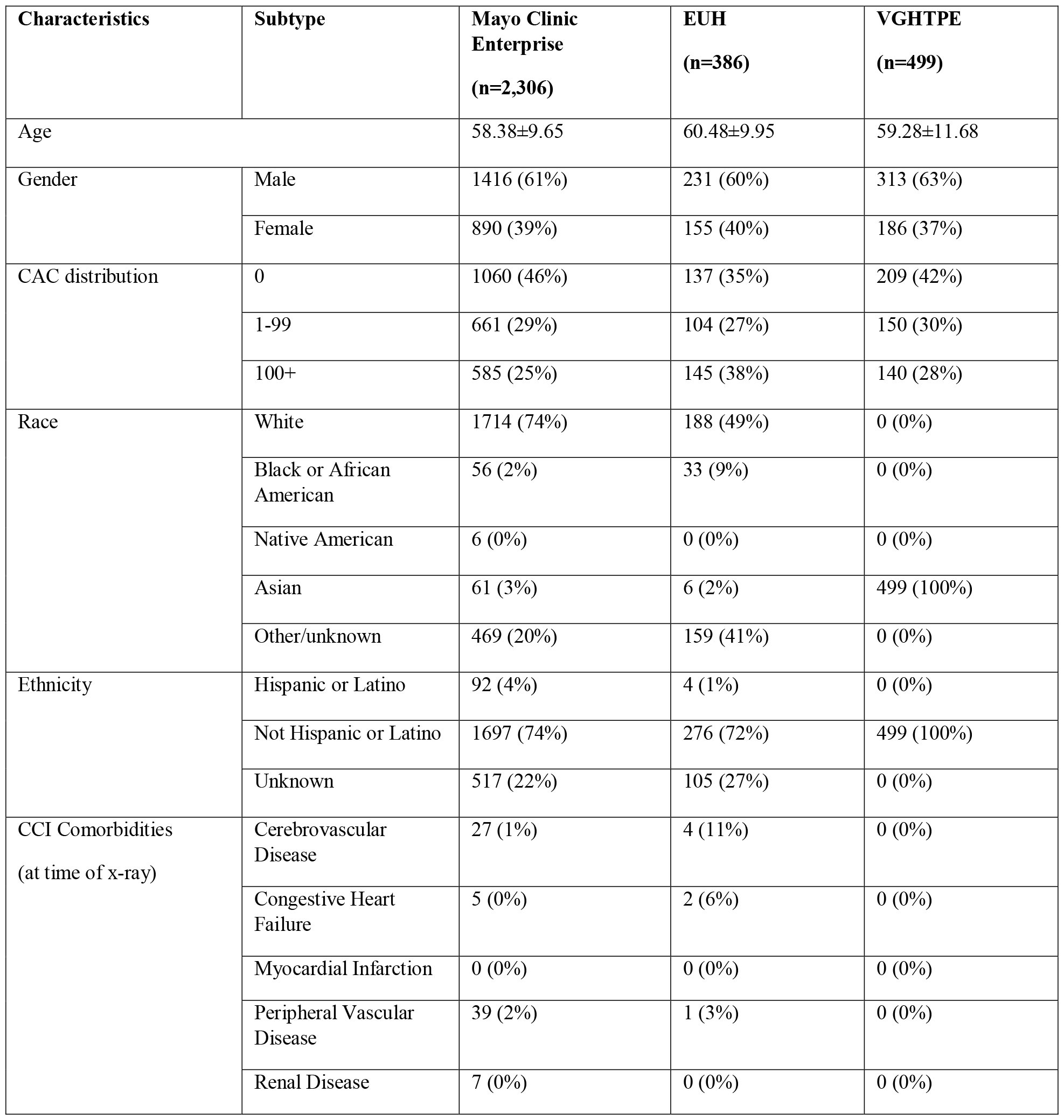
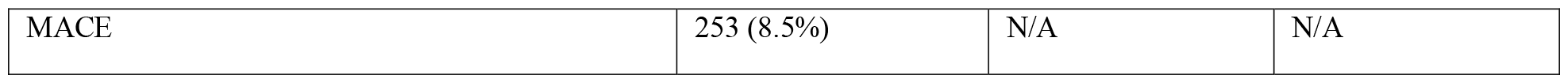
Demonstrate the patient characteristics for internal and two external institutions (domestic and foreign). MACE outcomes were not available on the external dataset.

| Characteristics | Subtype | Mayo Clinic Enterprise<br>(n=2,306) | EUH<br>(n=386) | VGHTPE<br>(n=499) |
| --- | --- | --- | --- | --- |
| Age |  | 58.38±9.65 | 60.48±9.95 | 59.28±11.68 |
| Gender | Male | 1416 (61%) | 231 (60%) | 313 (63%) |
|  | Female | 890 (39%) | 155 (40%) | 186 (37%) |
| CAC distribution | 0 | 1060 (46%) | 137 (35%) | 209 (42%) |
|  | 1-99 | 661 (29%) | 104 (27%) | 150 (30%) |
|  | 100+ | 585 (25%) | 145 (38%) | 140 (28%) |
| Race | White | 1714 (74%) | 188 (49%) | 0 (0%) |
|  | Black or African American | 56 (2%) | 33 (9%) | 0 (0%) |
|  | Native American | 6 (0%) | 0 (0%) | 0 (0%) |
|  | Asian | 61 (3%) | 6 (2%) | 499 (100%) |
|  | Other/unknown | 469 (20%) | 159 (41%) | 0 (0%) |
| Ethnicity | Hispanic or Latino | 92 (4%) | 4 (1%) | 0 (0%) |
|  | Not Hispanic or Latino | 1697 (74%) | 276 (72%) | 499 (100%) |
|  | Unknown | 517 (22%) | 105 (27%) | 0 (0%) |
| CCI Comorbidities<br>(at time of x-ray) | Cerebrovascular Disease | 27 (1%) | 4 (11%) | 0 (0%) |
|  | Congestive Heart Failure | 5 (0%) | 2 (6%) | 0 (0%) |
|  | Myocardial Infarction | 0 (0%) | 0 (0%) | 0 (0%) |
|  | Peripheral Vascular Disease | 39 (2%) | 1 (3%) | 0 (0%) |
|  | Renal Disease | 7 (0%) | 0 (0%) | 0 (0%) |

|  |  |  |  |
| --- | --- | --- | --- |
| MACE | 253 (8.5%) | N/A | N/A |

### External cohort

To test the model’s robustness and generalization capabilities, we collected testing data from two external healthcare centers *(domestic and foreign)* - Emory University Healthcare (EUH), and Taipei Veterans General Hospital (VGHTPE approved by IRB: 2023-09-009CC) in Taipei, Taiwan, with significantly varying patient populations in terms of race and ethnicity. To match the internal data, we applied the same patient selection criteria - patients who had a coronary CT scan between 2012 - 2022 and a CXR exam with frontal view within a ±1 year period of the CT (Table 1). CAC scoring was performed using Agatston scoring methods (14). Only a limited number of randomly selected cases were used as testing due to legal regulation and the time-consuming de-identification process.

### Multi-channel image formation

All the images were converted from Digital Imaging and Communications in Medicine (DICOM) file format to JPEG images using the open-source Niffler (https://github.com/Emory-HITI/Niffler) library (17). During conversion, images were kept in the original, native high-resolution image and kept in 16-bit gray-scale. During initial experiments, the CNN-based imaging models were noted to focus on the shoulder and neck regions for potentially determining the bone density and age to predict CAC scores. To reduce the learning of spurious correlation and allow the image model to focus on the coronary arteries, we first used a pre-trained lung segmentation model (18) to segment the lungs and computed a tight bounding box for cropping the center chest area. Then, we inverted the lung mask to obtain a rough segmentation of the heart and generate a lung masked image. An off-the-shelf bone suppression code (18) was used to suppress the ribs in the chest x-ray images to hide the bone density (19). Finally, we combined the original cropped chest centered image, lung masked image and rib bone suppressed image to generate a three-channel image from each frontal CXR image (see Fig 1).

**Figure 1.**
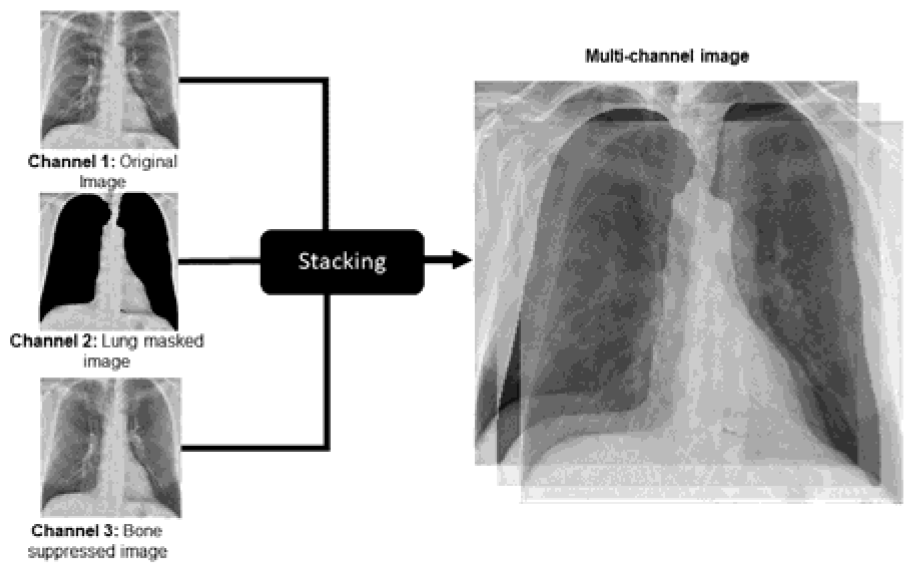
Multi-channel image formation – formed by stacking the original cropped chest centered image, lung masked image, and rib bone suppressed image.

### Multitask Image Classification Model

Given the complexity of the CAC detection task, we designed a multitask learning (MTL) paradigm by combining the MACE (major adverse cardiovascular event) prediction task (20) which includes acute myocardial infarction, stroke, hospitalization due to cardiac event, and cardiovascular mortality and is ideally related with the CAC detection. This parallel task can help extract additional information to support the primary CAC prediction. Based on chart-review, we manually curated MACE events within 2 years of the CXR study. We design the MTL paradigm with joint learning where both tasks are optimized with weighted loss: *l*_*total*_ = *γ*_1_*l*_*MACE*_ + *γ*_2_*l*_*CAC*_, where *γ* is the weighting parameter and *l*_*MACE*_, *l*_*CAC*_ are the individual branch losses (Fig 2). We trained a ResNeXt101 (21) backbone with the MTL strategy that uses shared CNN for both tasks and the backbone will be updated with the loss from both tasks. This strategy would have a synergistic effect on the backbone training (22).

**Figure 2.**
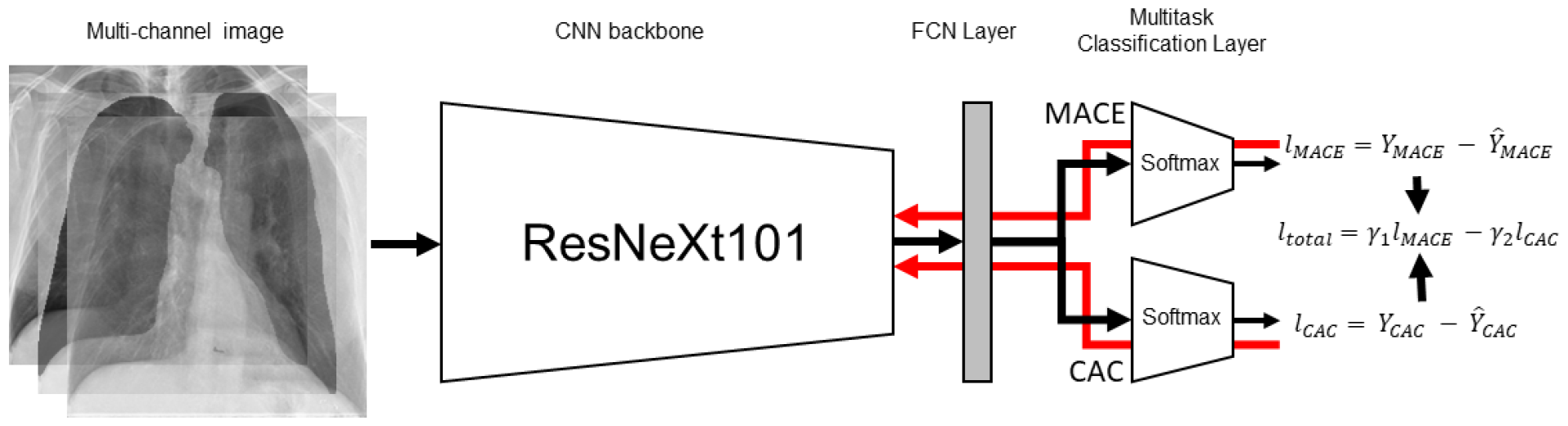
Multitask model. A ResNeXt101 backbone was used to classify MACE event and CAC category in parallel with weighted loss.

### Fusion Model

To provide additional data about patient and acquisition protocol which is easy to obtain during the CXR imaging, we combined simple tabular data - patient demographics (age, gender) and X-ray manufacturer category with the CXR image using late fusion strategy (decision level fusion) (23). Addition of patient demographics and device information may also help to reduce the bias in the model and allow generalization. We trained an individual supervised model (random forest) for the tabular data and a meta learner model that takes input CAC task prediction probability from both the MTL image model and tabular model and creates an aggregated function of the probabilities. With optimal learnt weight by the meta-learner, theoretically, better or equal performance can be achieved compared to either of the individual modality models.

## Results

We evaluated the MTL CAC fusion model on a hold-out test dataset from Mayo clinic (n=341) and independent dataset from EUH (n=386) and VGHTPE (n=499) using standard statistical metrics - precision, recall, and f1-score (Table 1). The optimal operating point was selected from the receiver operating characteristic curve (ROC) and in Fig. 3, we reported the class-wise area under the ROC (AUROC) using a one-vs-all strategy to assess the model’s probabilistic diagnostic accuracy. On the internal testset, the model achieved 0.72 and 0.66 AUROC for the ≥100 CAC category (clinically significant CAC) and 0 CAC category respectively. The performance was suboptimal for the intermediate 0-99 CAC category (0.58 AUROC), with similar trend observed for the EUH and VGHTPE external datasets. For the overall three class CAC detection, the performance remained moderate with average f1-score - *0*.*66 for Mayo, 0*.*62 for EUH and 0*.*61 for VGHTPE cohorts*. Despite a wide racial and ethnic difference between the centers, the performance remained consistent across the external setting.

**Figure 3.**
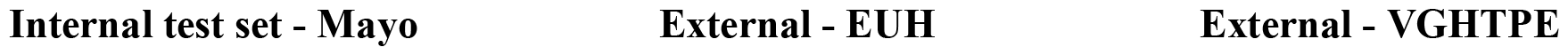
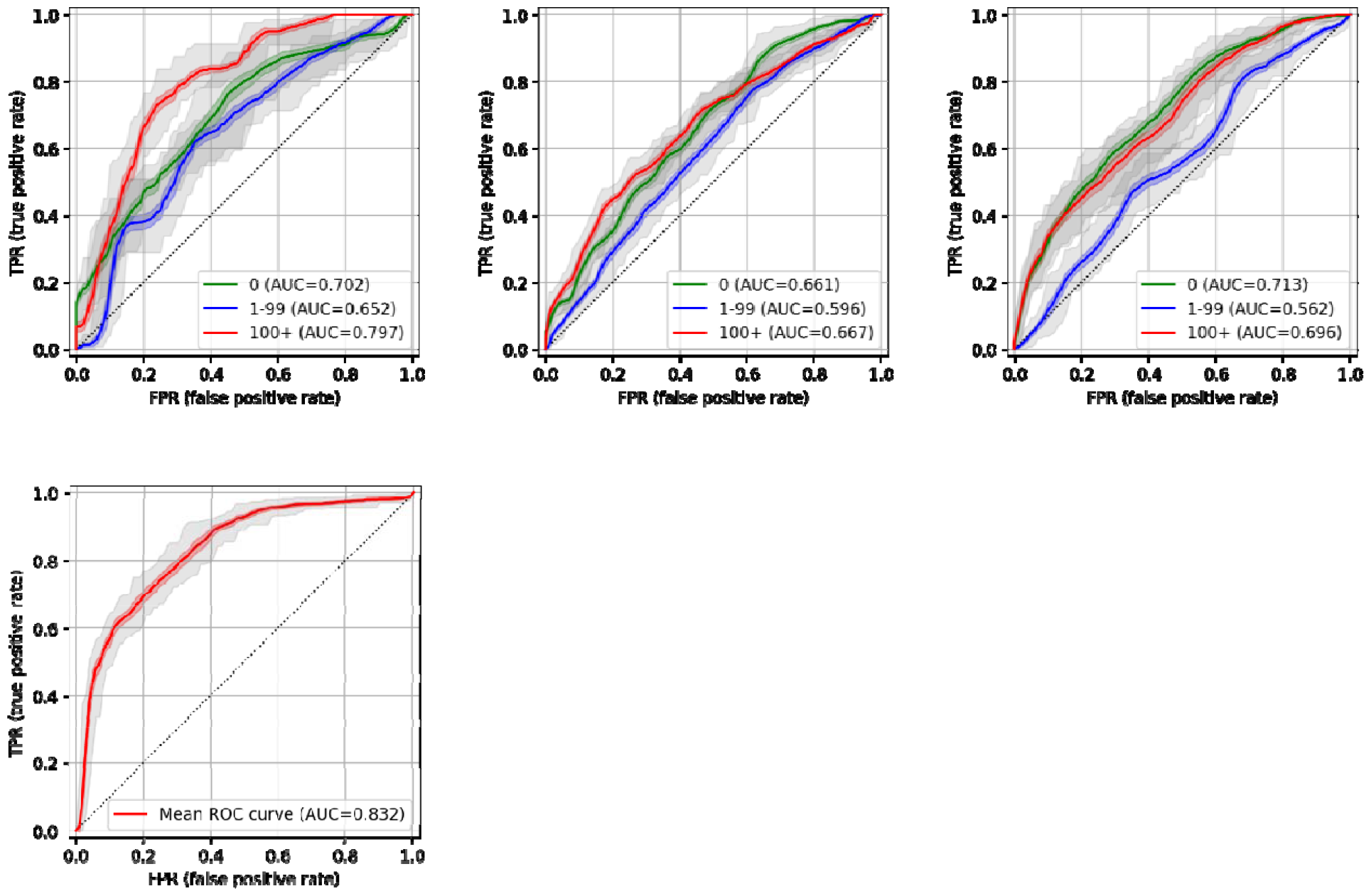
Top row: MTL fusion model Receiver Operating Characteristic (ROC) curve for discrimination of CAC category on the Mayo Internal hold-out testset, external EUH, and external VGHTPE. Shaded regions display 95% confidence interval; bottom row: MTL fusion model Receiver Operating Characteristic (ROC) curve for discrimination of MACE on the Mayo Internal hold-out testset.

### Internal test set - Mayo External - EUH External - VGHTPE

MACE prediction was performed as a *parallel auxiliary task within the MTL paradigm* and only evaluated on the Mayo holdout test set since the MACE outcome was not available on the external EUH and VGTHPE datasets (Fig. 3). The model achieved 0.83 AUROC score, and 0.954±0.023 precision and 0.828±0.019 recall for identifying the MACE category.

#### Discrimination of ‘high’ and ‘low’ CAC group

Although detection of 100+ CAC score from cardiac CT is important for clinical intervention with a statin, opportunistic screening for the detection of higher risk CAC from regular low cost CXR imaging is also clinically useful to triage patients to further dedicated diagnostic evaluation. Thus, we evaluate the MTL fusion model performance for binary classification tasks to differentiate high CAC candidates (Fig 4) and we experimented with both ‘0 vs 100+’ and ‘0 vs 400+’. The model demonstrated high performance (0.84 average AUROC) for differentiating high from low plaque deposits using only CXR imaging and achieved fair performance (0.75 average AUROC) for moderate plaque on both internal and external datasets. This suggests this MTL fusion model may identify the 400+ category with 84% confidence from the regular CXR imaging.

**Figure 4.**
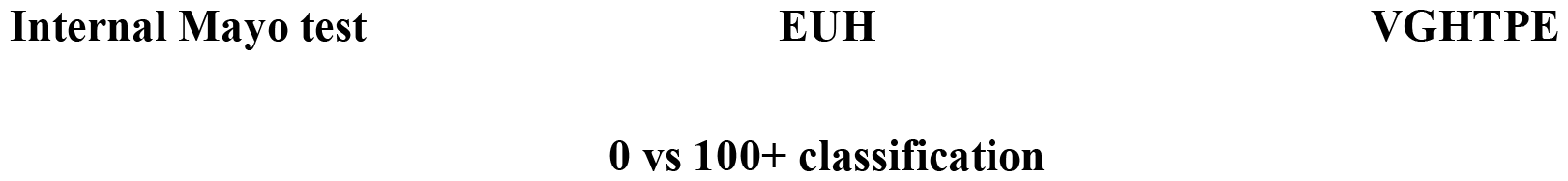
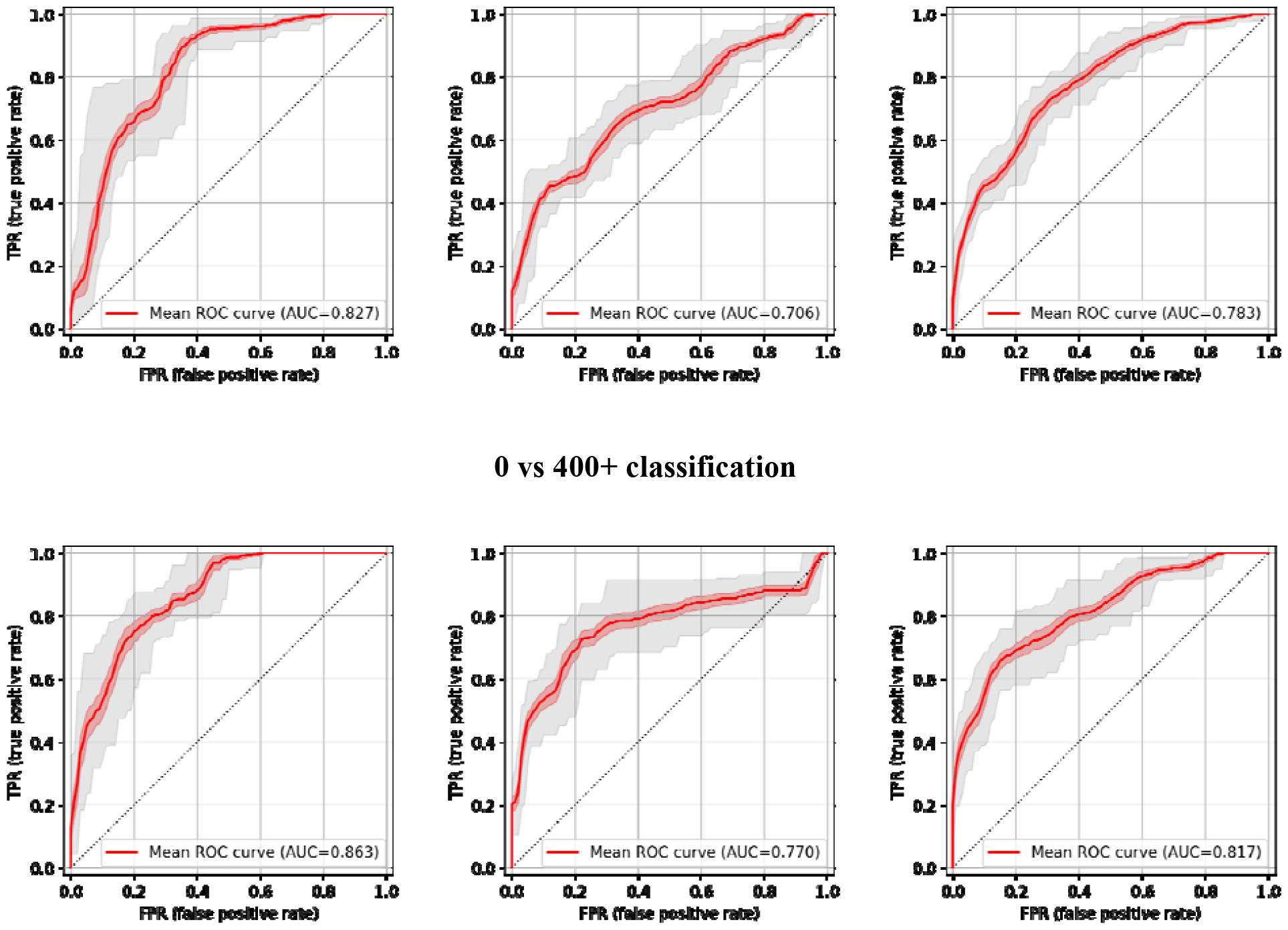
MTL fusion model Receiver Operating Characteristic (ROC) curve for discrimination of ‘high’ and ‘low’ CAC group on the Mayo Internal hold-out testset and EUH and VGHTPE external datasets. Shaded regions display 95% confidence interval;

##### Qualitative interpretation

To better understand the potential limitations of our model and to improve its interpretability, a visual analysis of the features extracted from CXR in misclassified cases was performed using GRADCAM++ (24) (red-more important, blue - less important). We found that common scenarios in misclassified cases were the presence of cardiac devices, pleuropulmonary diseases (including pulmonary nodules and pleural diseases), and aortopathy (including tortuosity and calcification of thoracic aorta) (see Figure 6). In several false positive cases (meaning cases classified as CAC score >0 by the model when true CAC score was 0) the presence of pulmonary nodules and external devices were evident in the CXR and seemed to capture the model’s attention. In panel A nodular opacities are evident in the CXR and seemed to be captured by the model mainly in the right lung; external electrodes and cables are also present in this case. In the CXR depicted in panel B, numerous pulmonary nodules and tortuosity in the descending aorta seem to be captured by the model. Panel C provides additional evidence regarding how the model can recognize and capture external devices such as electrodes and cables. For several false negative cases (meaning cases misclassified 0 by the model when true CAC score was 100+), pleuropulmonary diseases were evident. In panel E, and in panel D to a lesser extent, the model seems to focus on the prominent pulmonary vasculature of these cases. Additionally, in panel E mild calcification of thoracic aorta can be noticed. From a clinical perspective, it was difficult to identify definitive causes or imagining findings that could generate misclassifications by the model. However, after performing this qualitative interpretation, we could suggest that the model should be used with more caution in patients with pleuropulmonary diseases, external devices and aortopathy. Further investigation in the explainable artificial intelligence field will be critical to confirm our findings and to secure that models involving cardiac imaging are more understandable to future users.

**Fig. 6.**
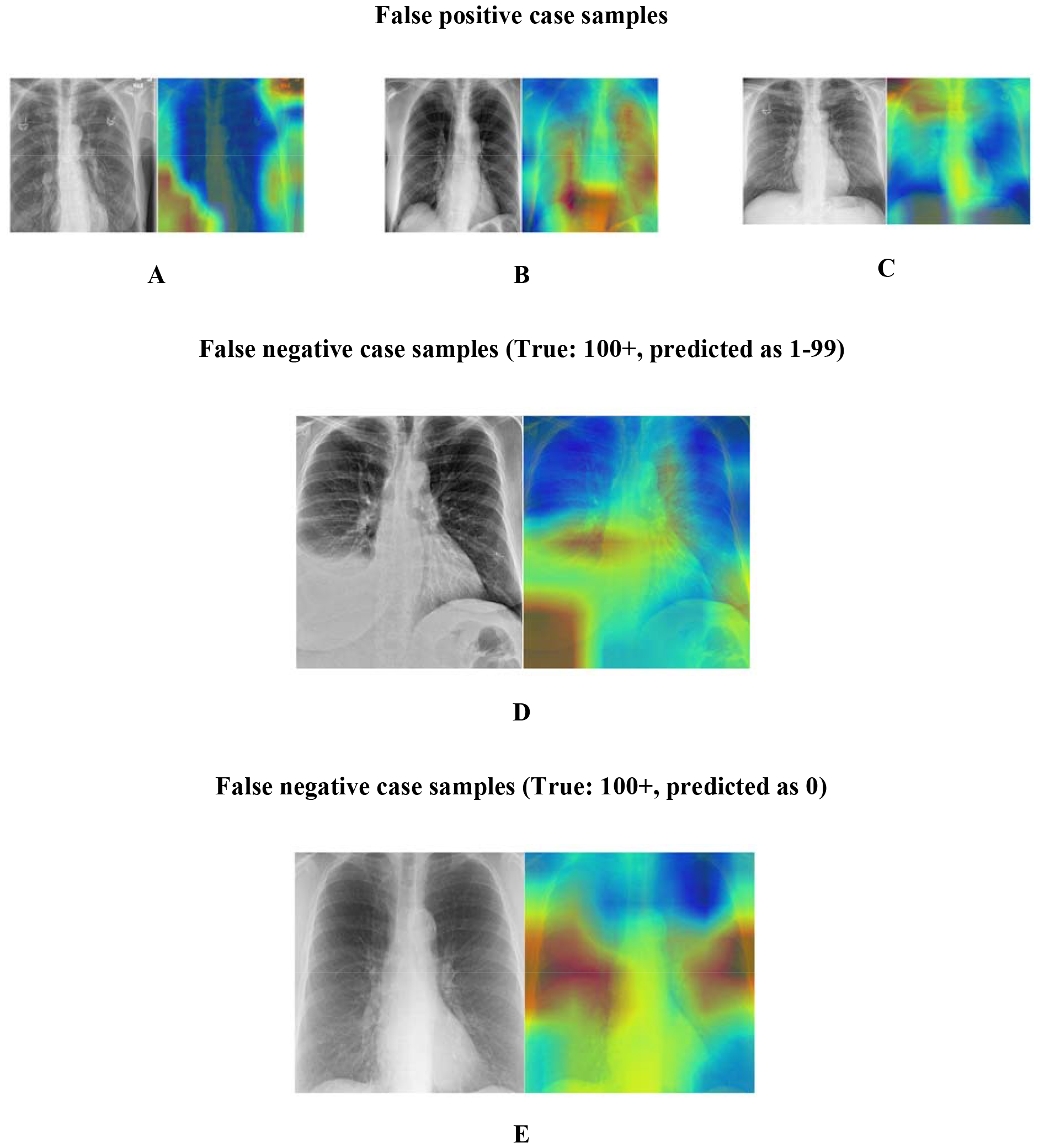
Model interpretation of false positive and negative case using GADCAM++.

## Discussion

The major contribution of this study is the development and external validation of an CXR-based AI model for opportunistic CAC screening and identify patients with undiagnosed coronary atherosclerosis. This approach utilizes a multitask fusion learning model to extract calcification features from routine CXRs and classify CAC scores, with external validation on datasets containing significantly different population representations (Mayo internal 74% white, EUH 50% African American, and VGTHPE 100% Asian). Additionally, incorporating clinical factors such as age, sex, and frailty score further enhanced the model performance in CAC classification. Although the model performance was moderate on the three CAC group classification (0 vs 1-99 vs 100+) with 0.65 AUROC on Mayo test and average 0.57 AUROC on VGTHPE and EUH, the model achieved high performance (0.84 average AUROC) for differentiating high (400+) from 0 CAC score which is the primary task of the opportunistic screening model. Trained model and the inference code can be downloaded with an academic open-source license from https://github.com/jeong-jasonji/MTL_CAC_classification.

### Enhancing model performance with clinical information

Patient age, sex, frailty score, and vendor information have been identified as significant contributors to the CXR-CAC model’s predictive capabilities. This observation implied the correlations between CAC scores and these patient factors, which is in line with the recent recommendations on CAC score interpretation (25). The inclusion of some clinical information alongside the visual features extracted from CXR images enhances the model’s ability to estimate CAC scores accurately and offers a more holistic approach to cardiovascular risk assessment.

### Multitask learning (MTL) paradigm with MACE prediction as auxiliary task

We addressed the complexity of the CAC prediction task from CXR by designing a multitask learning paradigm where MACE prediction is coded as an auxiliary task given the intuition that the CAC score should be highly correlated with MACE. The proposed model can simultaneously predict CAC category and MACE from the CXR, and the model achieved high performance for MACE prediction on the internal dataset. Using an ablation study, the details of which are given in the supplementary materials, we showed that the MTL model outperformed the single task learning model by a significant margin (0.58 to 0.65 AUROC). However, given complexity of the MACE outcome curation, we only validated the MACE branch on the internal dataset.

### Existing literature and comparative analysis

Recently, imaging modalities other than CT have been considered as potential alternatives for the assessment of CAC scores and its prognostic value with the assistance of AI models (9). Kamel et. al. (26) created a deep learning model to predict binary CAC classification (high versus low) from CXR images and reported 0.74 AUC (100+ CAC vs. 0 CAC) on frontal CXR and 0.7 (400+ CAC vs. 0 CAC) on lateral CXR images, however only validated on a single institutional data, so its generalizability and clinical impact remains unclear (27). Interestingly, their model had worse performance in handling cases with more distinct CAC scores. Our AI-CXR model exhibited high discriminatory abilities (0.83-0.86 AUROC) in predicting zero CAC and 400+ CAC groups and the findings were confirmed on external datasets.

Yuan et al. (9) reported a video-based artificial intelligence (AI) convolutional neural network which was trained to predict zero versus high (400+) CAC scores from parasternal echocardiography. While demonstrating good performance (0 CAC: 0.81 and 400+ CAC: 0.74), the study used a relatively clear cutoff and did not address patients with intermediate-risk (CAC score between 0 to 400), which is a group with more challenges in preventive intervention decisions. Our model had similar performance in differentiating cases of CAC 0 and CAC 100+ (Figure 4); we believe the superior performance relates to the fundamental difference of the 2 modalities, in which x-ray covers the whole heart while TTE only provides slices through specific cardiac axes; additional x-rays are superior for the detection of calcification compared to ultrasound. Furthermore, while TTE is a radiation-free modality, its cost-effectiveness, turnaround time, and availability may not be as favorable as CXR in resource-limited practice scenarios. Finally, risk stratification based on TTE-predicted CAC showed similar prognostic value to CT CAC scores in predicting significant differences in 1-year survival rates among high-CAC patients. The Yuan et al. (9) study suggests that deep learning of TTEs holds promise for adjunctive coronary artery disease risk stratification and guiding preventive therapies.

### An AI-enabled opportunistic screening tool - broad impact

The CXR-CAC model has dual advantages that make it highly beneficial in the field of cardiovascular risk assessment. Firstly, CXR imaging is widely available and accessible across various healthcare settings, including primary care clinics, urgent care centers, hospitals, and even remote or resource-constrained areas (13,28). This widespread availability enables the model to facilitate opportunistic screenings, allowing for the identification of individuals at risk for cardiovascular disease without the need for specialized cardiac evaluations. This inclusive approach ensures that even patients who may not have access to cardiology specialists or awareness of cardiovascular screening can benefit from risk assessment using CXR imaging, particularly asymptomatic and young patients. *Without any additional co*st, w*e can also reuse the retrospective CXR ima*ges to stratify the wider population based on CAD risk.

Secondly, the CXR-CAC model offers the advantage of low radiation dose, making it suitable for repetitive screenings. In comparison to CT scans used for coronary artery calcification (CAC) assessment, CXR-based screening involves significantly reduced radiation exposure. The low radiation dose associated with CXR allows for repeated screenings over time, facilitating longitudinal tracking of cardiac health and the timely detection of potential risk factors. Healthcare providers can implement more frequent screenings using CXR as part of preventive care strategies, enabling closer monitoring of changes in cardiac health status and supporting early interventions and preventive measures.

The combination of CXR’s wide availability and low radiation dose, coupled with the power of AI in the CXR-CAC model, provides a simple, cost-effective, and efficient screening tool. By leveraging existing infrastructure and the widespread availability of CXR imaging, the CXR-CAC model enables targeted interventions, public health initiatives, and timely risk stratification for individuals at risk of cardiovascular disease in underserved areas. This has the potential to improve patient outcomes and contribute to the early detection and prevention of cardiovascular conditions on a population scale. In areas where resources, infrastructure, or expertise for cardiac CT are scarce, this approach has the potential to broaden the availability of CVD screening and risk stratification, thereby enabling timely interventions and preventive measures for individuals in underserved areas.

#### Limitations

The study only considers the frontal view of the CXR image due to wider availability. In future, lateral view can also be assessed as it may provide additional data to boost the model. The model only obtained moderate performance for the three class CAC detection task; however, given the opportunistic screening goal of the framework, the binary risk stratification model obtained an impressive 0.84 average AUROC score on the internal and external validation cohort and should be capable in differential screening. Given training with the MTL paradigm with MACE, the model derived misclassification cases for pleuropulmonary conditions and aortic calcification. There was some bias of false negative rates across different subgroups (gender, race, and age) that we included in the supplementary materials that will need to be address in future studies. Additionally, we did not have BMI information at the time of the study to include in our bias analysis as a higher BMI is associated with higher risk of CAC (29). There was also some sampling bias in our dataset where we generally had a higher number of patients with pulmonary nodules with low CAC score. Further research and validation are warranted to optimize the model’s performance and evaluate its real-world clinical utility.

## Data Availability

The data is internal and is not available for access at the moment.

