## Supplemental materials for "Opportunistic screening for coronary artery calcium deposition using chest radiographs – a multi-objective models with multi-modal data fusion"

**Supplementary document**

*Table S1. MTL Fusion model internal and external test results for CAC in-terms of precision, recall and f1-score. 95% Confidence interval is calculated using auto-bootstrapping.*

| **CAC Category** | **Mayo Internal Test** | | | **External EUH** | | | **External VGHTPE** | | |
| --- | --- | --- | --- | --- | --- | --- | --- | --- | --- |
|  | **Precision** | **Recall** | **F1-score** | **Precision** | **Recall** | **F1-score** | **Precision** | **Recall** | **F1-score** |
| 0 | 0.748±0.037 | 0.724±0.038 | 0.729±0.036 | 0.758±0.027 | 0.749±0.032 | 0.746±0.033 | 0.680±0.047 | 0.645±0.018 | 0.656±0.025 |
| 1-99 | 0.627±0.062 | 0.550±0.040 | 0.565±0.037 | 0.555±0.079 | 0.434±0.022 | 0.458±0.022 | 0.602±0.068 | 0.494±0.020 | 0.514±0.015 |
| 100+ | 0.780±0.052 | 0.653±0.028 | 0.683±0.024 | 0.700±0.059 | 0.623±0.020 | 0.645±0.026 | 0.683±0.048 | 0.637±0.013 | 0.650±0.020 |
| Overall | 0.764±0.048 | 0.687±0.047 | 0.705±0.037 | 0.682±0.047 | 0.642±0.016 | 0.654±0.023 | 0.730±0.055 | 0.686±0.069 | 0.696±0.059 |

*Table S2. MTL Fusion model internal test results for MACE in-terms of precision, recall and f1-score. 95% Confidence interval is calculated using auto-bootstrapping.*

| **MACE Category** | **Mayo Internal Test** | | |
| --- | --- | --- | --- |
|  | **Precision** | **Recall** | **F1-score** |
| No MACE | 0.916±0.044 | 0.640±0.024 | 0.747±0.027 |
| MACE | 0.954±0.022 | 0.828±0.019 | 0.877±0.017 |
| Overall | 0.935±0.040 | 0.734±0.097 | 0.812±0.069 |

***Ablation Studies:*** We performed ablation studies to evaluate - (i) the relevance of multitask learning paradigm and (ii) inclusion of multimodal data (image + tabular) for the opportunist screening. For the first ablation, we compared the single task imaging model with only a single branch for CAC prediction against the MTL paradigm that was designed with parallel MACE and CAC prediction branches and presented the ROC curves in Fig 7 (single task) and Fig 3 & 4 (MTL). The single task model achieved 0.58 average AUROC on the internal Mayo test, while the MTL paradigm improved the performance up to 0.65. On the external EUH and VGHTPE test, the single task model average AUROC scores were 0.56 and 0.55 respectively, while the MTL model achieved 0.57 average AUROC for both EUH and VGHTPE. We also evaluated single tasks for the binary classification tasks (0 vs 100+, 0 vs 400+) on all the sites and the MTL model (averaged AUROC 0.82) performed consistently better than the single task (averaged AUROC 0.78).

For the *second ablation*, we compared the performance of the image only and EHR only model against the fusion model to demonstrate the power of combining image data with EHR. As seen from Table 2, EHR model outperformed image only model on the internal dataset (Mayo) but on the external dataset image only model outperformed EHR only model for the EUH testset which could be due to racial variations. However, the fusion model consistently outperformed both image only and EHR only model on all the test-sets.

*Table S3. Ablation study: comparative analysis using image-only, EHR-only and Fusion model. Optimal performance on the binary task (0vs100+) is highlighted in* ***bold****.*

|  | ***Image only model*** | | | ***EHR only model*** | | | ***Fusion model*** | | |
| --- | --- | --- | --- | --- | --- | --- | --- | --- | --- |
|  | **Mayo** | **EUH** | **VGHTPE** | **Mayo** | **EUH** | **VGHTPE** | **Mayo** | **EUH** | **VGHTPE** |
| *Precision* | 0.702±0.059 | 0.639±0.057 | 0.653±0.063 | 0.726±0.076 | 0.635±0.105 | 0.697±0.050 | **0.764±0.048** | **0.682±0.047** | **0.730±0.055** |
| *Recall* | 0.628±0.035 | 0.566±0.020 | 0.573±0.078 | 0.671±0.044 | 0.562±0.141 | 0.647±0.041 | **0.687±0.047** | **0.642±0.016** | **0.686±0.069** |
| *f1-score* | 0.648±0.030 | 0.583±0.021 | 0.591±0.067 | 0.686±0.051 | 0.570±0.13 | 0.661±0.036 | **0.705±0.037** | **0.654±0.023** | **0.696±0.059** |

***Bias analysis:*** We performed a bias analysis to evaluate if there was any disparity in false negative rates (FNR) in our MTL model across patient characteristics specifically age, gender, and race. We chose to focus on checking for bias in FNR, since the proposed opportunistic screening tool should minimize the false negative rate, in other word will not miss any high risk patients. Saleiro et al. defines disparity as a ratio between the metric of a group over the metric of a reference (majority) group (25). In each bias study, the study population was grouped according to their characteristics and bias disparity was calculated while keeping in mind the fairness standard of 1.0±0.2 [0.8, 1.2] where any ratio above 1.2 and below 0.8 are considered as an unfair and biased disparity. From our bias analysis, we could see that age and gender were the demographics in which we saw the most unfair disparities. We could see that younger patients (<40 yrs) had up to twice as much false negatives than the reference group (40-60 yrs) for higher CAC group, and. interestingly female patients also had up to twice as much false negatives than male patients for CAC 0 and 100+. This disparity could be due to the minimal representation of younger age group in our cohort and unbalance representation of female in the CAC categories. However, we did not observe any significant disparities in FNR for race across CAC groups, though white patients outnumbered other racial groups in the training dataset.

| **CAC: 0** | **CAC: 1-99** | **CAC: 100+** |
| --- | --- | --- |
| **FNR bias over patient age groups** | | |
| 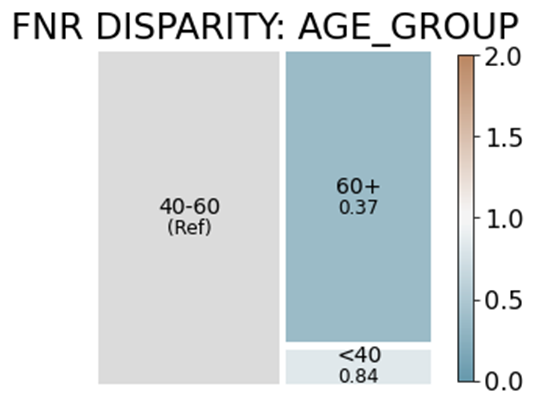 | 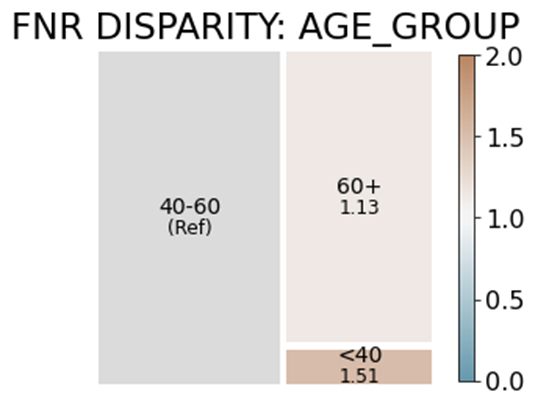 | 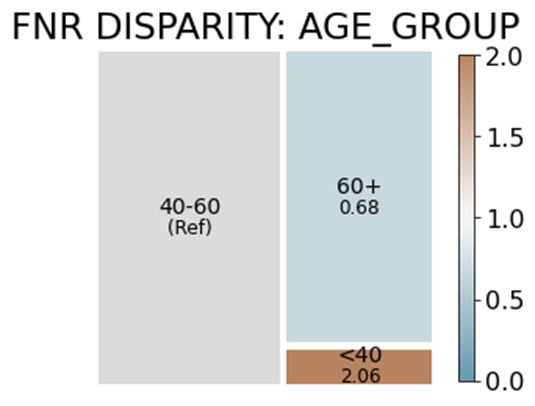 |
| **FNR bias over patient gender** | | |
| 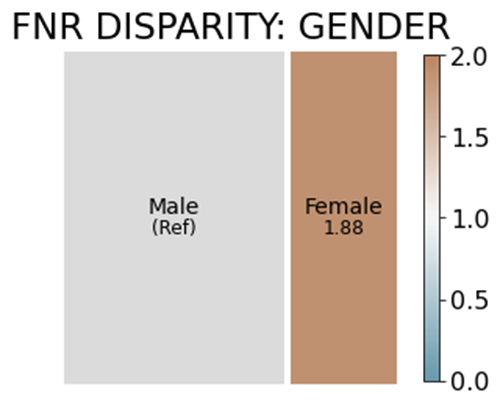 | 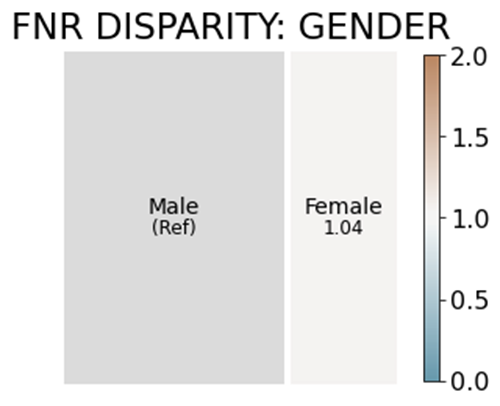 | 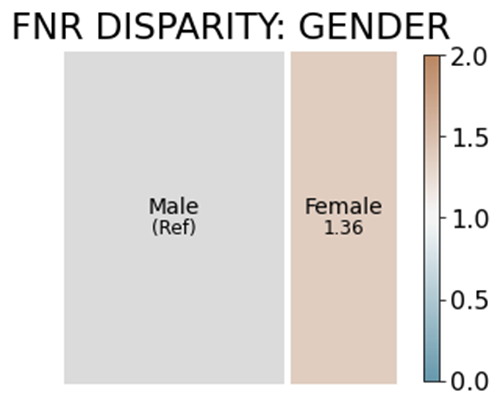 |
| **FNR bias over patient race** | | |
| 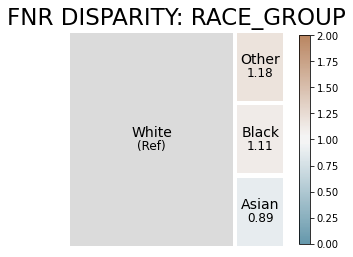 | 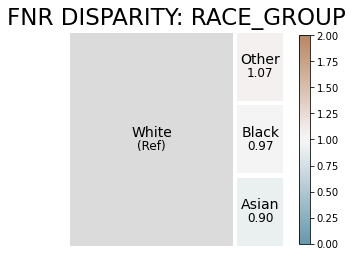 | 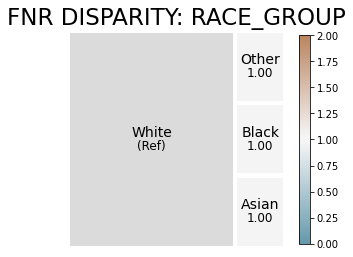 |

*Figure S1. Tree-plot to show False negative rate (FNR) bias across patient demographics. Based on 80% rule, 0.8-1.2 is considered to be fair disparity ratio.*
